# Biallelic variants in *HMGCS1* are a novel cause of rare rigid spine syndrome

**DOI:** 10.1101/2023.10.25.23297129

**Authors:** Lein NH Dofash, Lee B Miles, Yoshihiko Saito, Eloy Rivas, Vanessa Calcinotto, Sara Oveissi, Rita J Serrano, Rachel Templin, Georg Ramm, Alison Rodger, Joel Haywood, Evan Ingley, Joshua S Clayton, Rhonda L Taylor, Chiara L Folland, David Groth, Daniella H Hock, David A Stroud, Svetlana Gorokhova, Sandra Donkervoort, Carsten G Bönnemann, Malika Sud, Grace VanNoy, Brian E Mangilog, Lynn Pais, Marcos Madruga-Garrido, Marcello Scala, Chiara Fiorillo, Serena Baratto, Monica Traverso, Claudio Bruno, Federico Zara, Carmen Paradas, Katsuhisa Ogata, Ichizo Nishino, Nigel G Laing, Robert J Bryson-Richardson, Macarena Cabrera-Serrano, Gianina Ravenscroft

## Abstract

Rigid spine syndrome is a rare childhood-onset myopathy characterised by slowly progressive or non-progressive scoliosis, neck and spine contractures, hypotonia, and respiratory insufficiency. Biallelic variants in *SELENON* account for most cases of rigid spine syndrome, however, the underlying genetic cause in some patients remains unexplained.

In this study, we used exome and genome sequencing to investigate the genetic basis of rigid spine syndrome in patients without a genetic diagnosis. In five patients from four unrelated families, we identified biallelic variants in *HMGCS1* (3-hydroxy-3-methylglutaryl-coenzyme A synthase). These included six missense variants and one frameshift variant distributed throughout *HMGCS1*. All patients presented with spinal rigidity primarily affecting the cervical and dorsolumbar regions, scoliosis, and respiratory insufficiency. Creatine kinase levels were variably elevated. The clinical course worsened with intercurrent disease or certain drugs in some patients; one patient died from respiratory failure following infection. Muscle biopsies revealed irregularities in oxidative enzyme staining with occasional internal nuclei and rimmed vacuoles.

*HMGCS1* encodes a key enzyme of the mevalonate pathway, disturbance of which is also associated with *HMGCR*-limb girdle muscular dystrophy and *GGPS1*-muscular dystrophy. Quantitative PCR and western blotting confirmed HMGCS1 abundance in skeletal muscle and myogenic precursors. HMGCS1 levels in skeletal muscle were comparable between healthy controls and the index case with a homozygous p.(S447P) substitution. Muscle RNA-seq for a patient with a frameshift variant (c.344_345del:p.S115Wfs*12) and an in trans substitution (p.(Q29L)) showed *HMGCS1* transcript levels reduced to 53% compared to controls. The substitution appeared homozygous on RNA-seq, suggesting the allele harbouring the frameshift variant undergoes nonsense mediated decay.

*hmgcs1*^*-/-*^ zebrafish displayed severe early defects, including immobility at 2 days and death by days 3 post-fertilisation. We anticipate that the variants observed in this cohort have subtle effects on HMGCS1 function given most patients survived to adulthood. In support of the variants being hypomorphic, analyses of recombinant human HMGCS1 protein and four mutants (p.S447P, p.Q29L, p.M70T, p.C268S) showed all mutants maintained their secondary structure and dimerized state and had enzymatic activity comparable to the wildtype. Thermal stability of the mutants was similar or slightly reduced compared to the wildtype. Altogether, our analyses suggest that the identified missense variants in HMGCS1 act through a hypomorphic mechanism yet to be elucidated.

Here, we report an additional component of the mevalonate pathway associated with myopathy and suggest biallelic variants in *HMGCS1* should be considered in patients presenting with an unresolved rigid spine phenotype.

## Introduction

Inherited myopathies are a clinically and genetically heterogeneous group of debilitating disorders characterised by skeletal muscle dysfunction and weakness.^1^ Traditional subgroups include congenital myopathies, muscular dystrophies, mitochondrial myopathies, and metabolic myopathies. However, the clinical overlap between these entities can make it difficult to attain an accurate diagnosis.^1,2^

Rigid spine syndrome is a distinctive manifestation seen in various myopathies.^3-5^ Key features include contractures of the limb and spinal joints, limitation of flexion of the neck and trunk, weakness of the cervical and dorsolumbar spine muscles, and progressive scoliosis.^3-10^ Muscle pathology is variable and can include multiminicores, central cores, rods, rimmed vacuoles, and fibre-type predominance.^3,4,11^ The syndrome typically manifests during the first decade of life. It may be severe, slowly progressive, or non-progressive and may be accompanied by elevated serum creatine kinase levels.^2^ Patients may be susceptible to recurrent infections and respiratory insufficiency which can be fatal.^3,6-10^ For the purposes of this manuscript, the term rigid spine syndrome will be used as an umbrella description for disorders associated with the rigid spine features described above.

The genetic mechanisms and pathways that underlie rigid spine syndrome are diverse.^5,7,10,14,15^ Biallelic variants in *SELENON* encoding selenoprotein N are the most frequently reported cause of rigid spine manifestations.^8,10,12^ Other associated genes include *RYR1*^16^, *TOR1AIP1*^7^ *TPM3*^17^, *ACTA1*^9^, *TTN*^*18*^, *FHL1*^*19*^, and more recently, *GGPS1*.^11^ However, the genetic cause in some patients remains unknown despite screening known disease-associated genes.^13^ This suggests that additional genes are yet to be associated with rigid spine syndrome.

In this study, we investigated the genetic cause of rigid spine syndrome in patients remaining without a genetic diagnosis, using exome or genome sequencing and functional genomic approaches. In five patients from four unrelated families, we identified biallelic variants in *HMGCS1*, a gene of the mevalonate pathway encoding 3-hydroxy-3-methylglutaryl-coenzyme A synthase. To our knowledge, *HMGCS1* has not been previously associated with disease, though overlapping myopathic manifestations have been associated with both acquired and inherited disorders of the mevalonate pathway.^11,20,21^ Here, we report biallelic variants in *HMGCS1* as a novel cause of rigid spine syndrome which we have termed *HMGCS1*-myopathy. This report expands the associations of the mevalonate pathway with inherited skeletal muscle disease and encourages screening of *HMGCS1* in patients with unsolved rigid spine myopathy.

## Materials and methods

All investigations were approved by the Human Research Ethics Committee of the recruiting centres (University of Western Australia Human Research Ethics Office, National Center of Neurology and Psychiatry in Japan Human Research Ethics Office, Gaslini Children’s Hospital (Comitato Etico della Regione Liguria (163/2018), and the Mass General Brigham Institutional Review Board (Protocol #:2016P001422)). All families participating in this study provided informed consent. Zebrafish experiments and maintenance were approved by Monash Animal Research Precinct 3 committee.

### Clinical investigations

#### Patient details

We have clinically characterised five patients from four unrelated families originating from European or Asian descent. Patients presented with early onset rigid spine syndrome. Serum CK levels were measured in all patients including during acute episodes following intercurrent disease in some patients. Patients were also evaluated for pulmonary function and cardiac involvement. Muscle MRI was performed in some cases. Genomic DNA was isolated from peripheral blood of the patients and their relatives, where available.

#### Muscle pathology

Muscle biopsies were obtained from some patients as part of routine diagnostic investigations. The samples were frozen and processed for routine histological and histochemical analyses as outlined previously.^22^

### Genetic investigations

#### SPA1

Whole exome sequencing (WES) was performed on the affected siblings (patients P1 and P2) and their unaffected parents at the Center for Mendelian Genomics (Broad Institute of Harvard and MIT, Boston, USA). Exome capture was accomplished using an Illumina 37 Mb exome target (150 bp paired reads). Variant curation was performed in seqr (https://seqr.broadinstitute.org/).^23^ Variants were filtered based on a presumed autosomal recessive mode of inheritance and were restricted to rare (frequency<0.01 in gnomAD) coding variants, including extended splice-site variants. The *HMGCS1* variant was confirmed by PCR and bi-directional Sanger sequencing (Australian Genome Research Facility (AGRF) Perth, Australia).

#### JPN1

After standard DNA extraction, WES was performed for patient P3, unaffected parents and two unaffected siblings at department of Neuromuscular Research (National Institute of Neuroscience in National Center of Neurology and Psychiatry, Tokyo, Japan). Samples were processed using TruSeq DNA PCR-free sample preparation and 150 bp paired-end sequencing was conducted on the NovaSeq 6000 platform (BGI, China). BWA (0.7.5a-r405, https://github.com/lh3/bwa) and Picard (1.141, https://github.com/broadinstitute/picard/issues/431) were used to map low-divergent sequences from the human genome reference (GRCh37). GATK 3.5 (https://gatk.broadinstitute.org/hc/en-us) was used to call variants. Variants were then annotated with ANNOVAR. The variants were filtered based on a presumed autosomal recessive mode of inheritance and was restricted to rare (frequency<0.01 in gnomAD) coding variants, including extended splice-site variants. Bi-allelic variants in *HMGCS1* were confirmed by Sanger sequencing of the parents.

#### ITA1

After standard DNA extraction, trio WES was performed for patient P4 and their unaffected parents as previously described.^24^ The selection of the coding regions and flanking intronic sequences was made using the enrichment kit IDT xGen Exome Research Panel v2 (34 Mb, 19.433 geni). The sequencing was performed using Illumina technologies (PE 2X150) with the platform Illumina Novaseq6000. The quality of the sequence reads was assessed through QC statistics with FastQC (http://www.bioinformatics.bbsrc.ac.uk/projects/fastqc). Reads were aligned to the reference human genome (hg38, UCSC assembly) using BWA with default parameters. The HaplotypeCaller algorithm within the Genome Analysis Toolkit (GATK) package was employed for quality score recalibration, indel realignment, and variant calling. Variants were then annotated with ANNOVAR. After filtering for variants with a minor allele frequency (MAF) ≤ 0.001 in gnomAD (https://gnomad.broadinstitute.org) and an in-house database of 6,500 exomes, the predicted impact of the candidate variants on protein structure and function was evaluated through *in silico* prediction tools.

#### USA1

Whole genome sequencing and data processing were performed by the Genomics Platform at the Broad Institute of MIT and Harvard. PCR-free preparation of sample DNA (350 ng input at >2 ng/ul) was conducted using Illumina HiSeq X Ten v2 chemistry. Libraries were sequenced to a mean target coverage of >30x. Genome sequencing data was processed through a pipeline based on Picard, using base quality score recalibration and local realignment at known indels. Reads were mapped to the hg38 using BWA aligner. Single nucleotide variants (SNVs) and insertions/deletions (indels) were jointly called across all samples using the GATK HaplotypeCaller package version 4.0. Default filters were applied to SNV and indel calls using the GATK Variant Quality Score Recalibration (VQSR) approach. Annotation was performed using Variant Effect Predictor (VEP). Lastly, the variant call set was uploaded to seqr for collaborative analysis between the Centers for Mendelian Genomics (CMG) and investigator.

### Analysis of the HMGCS1 substitutions

Positions of the *HMGCS1* substitutions were annotated on the *HMGCS1* gene (ENSG00000112972) and protein structure (PDB: 2P8U).^25^ The crystal structure for HMGCS1 was accessed from the Protein Data Bank (PDB; www.rcsb.org)^26^ and analysed using the PyMOL Molecular Graphics System, Version 2.4.1. Annotations including residues of the HMGCS1 active sites, CoA binding sites, and the salt bridge were obtained from the UniProt database (Q01581) and the ICn3D (I-see-in-3D) molecular structure viewer.^27^ For conservation analysis, HMGCS1 amino acid sequences were obtained from UniProt and aligned by Clustal omega alignment (https://www.ebi.ac.uk/Tools/msa/clustalo/).

### *HMGCS1* expression

#### Gene expression databases

Bulk RNA-seq data for *HMGCS1* (ENST00000325110) was obtained from the Genotype-Tissue Expression (GTEx) portal (https://gtexportal.org/) and NCBI (BioProject PRJEB4337). Cap Analysis Gene Expression (CAGE) data was obtained from the Functional Annotation of the Mammalian genome consortium (FANTOM5; https://fantom.gsc.riken.jp/5/).^28^

#### RNA sequencing

Skeletal muscle RNA-seq was performed for patient P3 (JPN1) and three unaffected control samples from the National Center of Neurology and Psychiatry. RNA was extracted with the RNeasy Plus Mini Kit (Qiagen, Valencia, CA, USA; 74136). Libraries were prepared from total RNA with a TruSeq Stranded Total RNA Library Prep Kit (polyA capture, Illumina), according to the manufacturer’s instructions, and 100 bp paired-end sequencing conducted on the NovaSeq 6000 platform; 8 Gb of sequence data encompassing *HMGCS1* were generated for each sample (Macrogen, Seoul, Korea). RNA-seq reads were mapped using Spliced Transcripts Alignments to Reference (STAR) software (https://github.com/alexdobin/STAR). The reads were processed using StrandNGS® v4.0 (http://www.strand-ngs.com/) and analysed by DESeq^29^ compared to the three unaffected control samples.

We used DROP v1.0.3^30^, as previously described^31^, to analyse aberrant gene expression amongst a cohort of 129 skeletal muscle RNA-seq from rare muscle disease patients and unaffected controls, including JPN1. DROP leverages OUTRIDER^32^, which uses a denoising autoencoder to control co-variation before fitting each gene over all samples via negative binomial distribution. Multiple testing correction was done across all genes per sample using DROP’s in-built Benjamini-Yekutieli’s false discovery rate (FDR) method.

#### Quantitative PCR

Quantitative real-time PCR (qPCR) was performed to investigate *HMGCS1* transcript levels in healthy human tissue and cell lines. RNA extraction, cDNA synthesis and qPCR were performed as previously described.^33^ Reactions (10 μL) were performed using 2X Rotor-Gene SYBR Green PCR master mix (Qiagen; 204076) and contained 1 μL diluted cDNA and 0.8 μM of forward and reverse primers (5’-TTCCCTTGCATCTGTTCTAGC-3’ and 5’-TTTTATCAAGAGCAGACCCCG-3’). Reactions were performed for biological replicates depending on availability of samples (*n* =2-7). Cycling was performed on a Rotor-Gene Q real-time PCR cycler (Qiagen) as follows: 95°C (5 min), 45 cycles of 95°C (10 s) and 60°C (15 s), and subsequent melt curve analysis (60-95°C; +1°C per cycle). Data were normalised using the delta Ct method^34^ by comparing to the geometric mean of two reference genes (*TBP* and *EEF2*).^35^

### HMGCS1 protein abundance

#### Western blotting

Protein extraction and western blotting were performed as previously described.^36^ Frozen cell pellets and shredded tissue were suspended in lysis buffer containing 2% SDS, 125 mM Tris (pH 6.8), and 1x PIC (Halt™ Protease Inhibitor Cocktail; Thermo Scientific) and homogenised by sonication. Protein concentrations were determined by a BCA protein assay (Pierce; #23227). Samples were prepared in loading buffer (10% SDS, 312.5 mM Tris (pH 6.8), 50% glycerol, bromophenol blue saturated solution, 50 mM DTT and 1x PIC). Extracts (12 μg) and recombinant HMGCS1 (5 ng; production described below) were separated on 4-12% Bis-Tris NuPAGE Novex polyacrylamide gels and transferred to PVDF transfer membranes (Thermo Fisher Scientific). Membranes were blocked with PBS + 0.1% Tween-20 and 5% skim milk for 1 h at room temperature (RT) and probed with an anti-HMGCS1 rabbit polyclonal antibody (MyBioSource; #MBS2026097; 1:800 incubated overnight at 4°C) and subsequently, a secondary goat anti-rabbit horseradish peroxidase antibody (Sigma; #A0545) for 1 h at RT. Membranes were imaged by chemiluminescent detection (Pierce ECL Plus kit; #32132) using the Invitrogen iBright FL1000 imaging system (Thermo Fisher Scientific). An anti-GAPDH antibody (Sigma; #G8795; 1:10,000) was used as a loading control. Relative HMGCS1 abundance was calculated using ImageJ.^37^

#### Identification of HMGCS1 in primary human myoblasts and myotubes by mass spectrometry

Myoblasts and myotubes (day 2 and day 8 of terminal differentiation) from three healthy human donors (18M [P01236-18M], 32F [P01034-32F] and 42F [P01269-42F]; Cook Myosite)^38^ were solubilised in 5% sodium dodecyl sulphate (SDS), 50mM tetraethylammonium bromide (TEAB) and sonicated with a probe sonicator for 10 s on/off cycles for 30 s on ice. Samples were quantified via bicinchoninic acid (BCA) protein assay (Thermo Fischer Scientific), normalised to 60 ug of total protein and prepared for liquid chromatography tandem mass spectrometry (LC-MS/MS) using micro S-trap™ columns (ProtiFi) as per manufacturer’s protocol. Protein reduction and alkylation were performed with 40 mM 2-chloroacetamide (CAA) and 10 mM tris(2-carboxyethyl)phosphine hydrochloride (TCEP) at 99°C for 5 min with 1500 rpm shaking. Proteins were digested at 1:25 trypsin:protein ratio overnight. LC-MS/MS data was acquired as previously.^39^ Raw data were processed using the Spectronaut platform (version 14, Biognosys) and searched against the UniProt human database (canonical peptides and isoforms, reviewed, 42,386 entries) and a peptide library containing 147,852 entries. Default settings were used with exception of single hit proteins were excluded from the data and precursors were filtered on Q-value with imputation strategy set to “none”. Number of peptides were plotted with GraphPad Prism 9.0.1 (La Jolla, USA).

### Functional genomic investigations

#### Recombinant protein expression

Bacterial pET-30a(+) expression constructs were synthesised by Genscript (https://www.genscript.com/). These encoded the coding DNA sequences for wildtype human HMGCS1 (ENST00000325110, HMGCS1^WT^), and mutants encoding one of the four missense variants: p.Ser447Pro (HMGCS1^S447P^), p.Gln29Leu (HMGCS1^Q29L^), p.Met70Thr (HMGCS1^M70T^), or p.Cys268Ser (HMGCS1^C268S^) identified in patients. The constructs were codon-optimised for *E. coli* expression and included a six-histidine tag at the N-terminus. Constructs were transformed into BL21-DE3 *E. coli* following a standard heat-shock method.^40^ Cultures were grown at 37°C in Luria-Bertani (LB) broth containing 35 μg/mL kanamycin. Expression was induced with 0.1 mM Isopropyl β-d-1-thiogalactopyranoside (IPTG) with overnight growth at 16°C. Protein extraction and purification were performed as previously described.^41^ Cultures were lysed by sonication in 50 mM Tris (pH 8.0), 250 mM sodium chloride, 10% v/v glycerol and 0.1% Triton X-100. The soluble fraction containing recombinant protein was incubated overnight at 4°C with resins (profinity IMAC, #156-0123) charged with 0.2 M Nickel (II) sulfate hexahydrate (Sigma). Proteins were eluted by washing in buffer containing 50 mM Tris (pH 8.0), 150 mM NaCl, and 5% glycerol and increasing concentrations of imidazole (0 to 300 mM). Eluants were concentrated to in buffer containing 50 mM Tris (pH 8.14), 150 mM NaCl and 5% glycerol by centrifuging at 3,000 g, 4°C using 30 kDa Amicon ultra centrifugal filter units (Sigma).

#### Size exclusion chromatography

Size exclusion chromatography was performed as previously described.^41^ Protein concentrates were loaded onto a XK16/600 Superdex 200 column (GE Healthcare Life Sciences) for purification. Columns were equilibrated in 150 mM NaCl, 50 mM Tris (pH 8.14), 10% v/v glycerol. Fractions were collected and tested for protein of the expected size by SDS-PAGE. Protein identity and purity were determined by mass spectrophotometry (Proteomics International, Perth). The concentration of recombinant protein was determined using a NanoDrop 2000 V1.5 (Thermo Scientific) at 280 nm. The exclusion coefficient and molecular weight of protein were calculated using the Expasy ProtParam webtool (https://web.expasy.org/protparam/).^42^ The extinction coefficient and molecular weight (excluding His-tags) for HMGCS1 was 65.39 mM^-1^cm^-1^ and 57.29 kDa, respectively.

#### Circular dichroism

For circular dichroism (CD) melting curve and spectra analyses, recombinant wildtype and mutant HMGCS1 were diluted to 0.1 mg/mL in 10 mM sodium phosphate buffer (pH 7.4). In technical replicates, 300 μL of protein and buffer were loaded into 0.1 cm quartz cuvettes and measured by a Jasco J-1500 CD spectropolarimeter. For the melting curves, samples were measured at temperatures ranging between 15−95°C at 220 nm and 229 nm at a rate of **+**0.25°C/min (30 s data integration time [DIT], 2.00 nm bandwidth). Throughout the melting curve process (approx. every 5°C), CD spectra were measured between 195−260 nm (0.2 nm data pitch, 4 s DIT, 2.00 nm bandwidth). Data were collected using the JASCO Spectra manager software (2.15.24). Cuvette baseline measurements were subtracted from the sample spectra measurements. Data were zeroed by subtracting the average signal between 250−260 nm. Melt curves were generated using GraphPad Prism 9.0.1 (La Jolla, USA) by fitting each dataset to a sigmoidal four parameter logistic regression curve. The fits generated for each melt curve were based on temperature ranges which best captured protein folding in that particular dataset. The half height value was taken as a measure of protein melting temperature (T_m_; temperature at which 50% of protein is denatured). Data were analysed by a one-way ANOVA followed by Dunnet’s multiple comparison test and paired t-test to measure statistical significance using GraphPad Prism 9.0.1 (La Jolla, USA).

#### Enzyme assays

HMGCS1 catalyses the condensation reaction between acetyl coenzyme A (Ac-CoA) and acetoacetyl coenzyme A (AcAc-CoA) to form 3-hydroxy-3-methylglutaryl coenzyme A (HMG-CoA).^43^ To investigate the effect of the missense substitutions on HMGCS1 activity, we adapted an assay described by Lowe and Tubbs (1985)^43^ which measures the decrease in absorbance (303 nm) as AcAc-CoA is consumed. The standard assay system contained 100 mM Tris/HCl (pH 8.0) and 40 mM MgCl_2_in a total volume of 200 μl. Reactions were performed in triplicate in a UV-star 96 well microplate (Greiner) at 25°C and initiated with 1 μg of enzyme or with buffer alone as a no enzyme control (NEC). Concentrations of Ac-CoA and AcAc-CoA ranged between 0-300 μM. The amount of AcAc-CoA consumed per reaction was determined in relation to a standard curve and reactions were fit to non-linear one phase decay plots using GraphPad Prism 9.0.1 (La Jolla, USA). The amount of AcAc-CoA consumed within the first 50 s of the reactions was taken as a measure of the initial velocity (V_0_).

### Generation of zebrafish hmgcs1 mutant line

The zebrafish hmgcs1 null mutant line was generated by injecting one-cell stage embryos with a mix containing two pre-complexed guideRNA’s (Integrated DNA Technologies, Alt-R™) targeting exon3 (GCTTGGTCAACGTACTGAGAtgg and GACACCACTAACGCCTGCTAtgg), Cas9 protein (HiFi v3 from Integrated DNA Technologies), 300 mM KCl, and 0.005% phenol red. Injected embryos were raised to adulthood and crossed to wildtype fish, and embryos screened for deletions using the following genotyping primers: hmgcs1_ex3_F CCATGTGGCCCAAAGATGTG, hmgcs1_ex3_R TGGACTCCACCCAGTTGAC. A founder passing on a 302 bp deletion in exon3 was identified.

### HMGCS1 rescue experiments

Human wildtype HMGCS1 was codon-optimised for zebrafish using CodonZ^44^ and synthesised with an Sp6 transcriptional start site and an *afp* 3’UTR by Genscript™. Capped mRNA for injection was synthesised using the mMessage mMachine Sp6 kit (Thermofisher Scientific). One-cell stage embryos from *hmgcs1* heterozygote incrosses were injected with a mix containing HMGCS1 mRNA at 18 ng/ul, 300 mM KCl, 0.005% Phenol red, and cascade blue labelled dextran (Molecular Probes). Control-injection mix did not contain mRNA. Embryo survival was recorded daily until 4 days post-fertilisation. Remaining embryos were genotyped to determine the number of homozygous mutants alive.

#### Electron microscopy

48 h post-fertilisation embryos were fixed in 0.1 M sodium cacodylate buffer containing 2% paraformaldehyde and 2.5% glutaraldehyde for 24 h and rinsed with 0.1 M sodium cacodylate buffer (3x 10 min). Embryos were genotyped prior to processing. Samples were processed using a method modified from Hua, Laserstein, Helmstaedter.^45^ All processing steps were aided by a microwave regime (Pelco Biowave) and performed at room temperature unless otherwise stated. Samples were post-fixed in 2% osmium tetroxide in 0.1 M sodium cacodylate buffer for 12 min (2 min on/off cycles, 150 W, under vacuum) which was reduced with 1.5% potassium ferrocyanide in 0.1M sodium cacodylate for 12 min (2 min on/off cycles, 150 W, under vacuum). Samples were thoroughly rinsed in MilliQ (mQ) H_2_O for 3 × 40 s (150 W, no vacuum). This was followed with 1% aqueous thiocarbohydrazide solution for 12 min at 50°C (2 min on/off cycles, 150 W, under vacuum) and again rinsed thoroughly in mQH_2_O (3x 40 s, 150 W, no vacuum). Samples underwent further osmification in 2% osmium tetroxide (aqueous) for 12 min (2 min on/off cycles, 150 W, under vacuum), at room temperature. After rinsing with mQH_2_O as previous, samples were exchanged into 1% uranyl acetate (aqueous) overnight at 4°C. The following day the temperature was increased to 50°C in the biowave for further processing for 12 min (2 min on/off cycles, 150 W, under vacuum). Samples were again rinsed with mQH_2_O as previous. The final staining step involved incubation in lead aspartate for 12 min at 50°C (2 min on/off cycles, 150 W, under vacuum) and rinsing in mQH_2_O (as previous). Samples were then dehydrated with increasing concentrations of ethanol before final dehydration with acetone (ethanol: 20%, 50%, 70%, 90%, 2 × 100%; acetone: 2 × 100%) for 40 s per step (150 W, no vacuum), and infiltrated with increasing concentrations of hard EPON resin in acetone for 3 min each step (25%, 50%, and 75%, 2 × 100%). Samples were left in fresh 100% resin overnight. After being transferred into a silicone mould, samples were polymerised into blocks in the oven at 60°C for 48 h.

Longitudinal sections were cut at 70 nm on a ultramicrotome (Leica UC7) and collected on copper mesh grids. Samples were imaged on a Fei Nova Nano SEM 450 using the STEM2 detector at 30 kV. Maps software package (Thermo Fisher) was used to automatically acquire tiled regions with a pixel size of 6 nm. Tiles were assembled using the deprecated Stitch Grid of Images plugin within Fiji.^46^

#### Mitochondrial quantification in electron microscopy images

The number of mitochondria within a randomly sampled somite were quantified in *hmgcs1*^+/+^ and *hmgcs1*^-/-^ using the Multipoint tool in Fiji.^46^ Abnormal mitochondria were scored as showing detachment or invagination of the inner membrane and expressed as the percentage of the number of abnormal mitochondria/total number of mitochondria. Somite area was quantified by drawing a rectangle around the somite using the polygon tool in Fiji and used to normalize the number of mitochondria for each sample. The researcher was blinded to the genotype and the order of images randomised, during mitochondria scoring and area measurement, with the genotypes revealed for statistical comparison conducted by another researcher.

### Data availability

The data and constructs generated by this study are available upon request.

## Results

### Clinical findings

We report five patients from four families presenting with a clinical picture of rigid spine syndrome (Fig 1, Table 1 and Supplementary material). Age-of-onset ranged from birth to childhood. There was no relevant family history of disease in any of the families. Features included spinal rigidity primarily affecting the cervical (*n*=5/5) and dorsolumbar (*n*=4/5) regions, and scoliosis (*n*=5/5). All patients had restrictive pulmonary function with a forced vital capacity ranging between 10-56%. Some patients also presented with weakness of the proximal (*n*=4/5) and distal (*n*=2/5) limbs, scapular winging (*n*=3/5), and bulbar weakness (*n*=1/5; Table 1). None of the patients presented with facial weakness, ophthalmoparesis, or ptosis. Baseline creatine kinase levels were variably elevated in all cases, ranging between 190-7890 IU/L (normal ∼70-170 IU/L^47^). In three patients (P1, P3, P5) episodes of muscle deterioration and respiratory crisis were triggered by intercurrent disease or certain drugs. Patient P2 (SPA1) died during their teenage years from such an episode after developing a respiratory infection followed by a rapid decline in muscle strength and respiratory function leading to respiratory failure. Patient P3 (JPN1) was also reported to experience such episodes and elevated creatine kinase levels during febrile illness. The episode of decline for patient P5 (USA1) was triggered after starting megestrol and slowly improved after discontinuing the drug.

**Figure 1.**
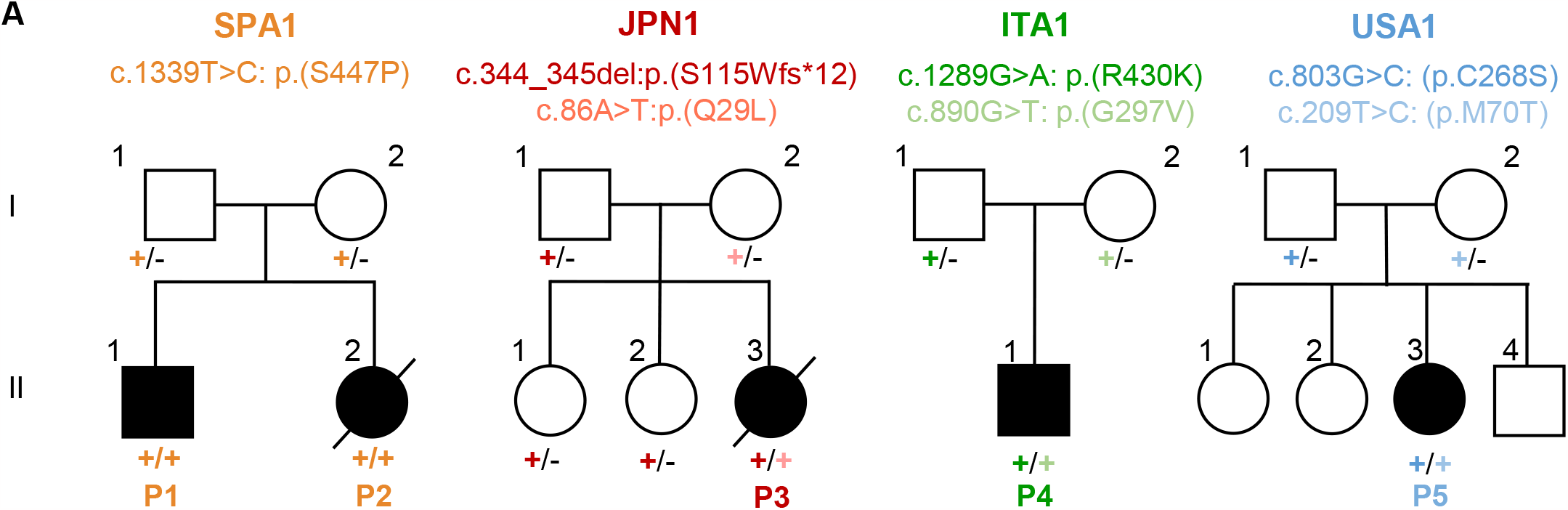
HMGCS1-myopathy cohort. **(A)** Pedigrees of the four families with biallelic variants in *HMGCS1* segregating with rigid spine syndrome.

**Table 1.**
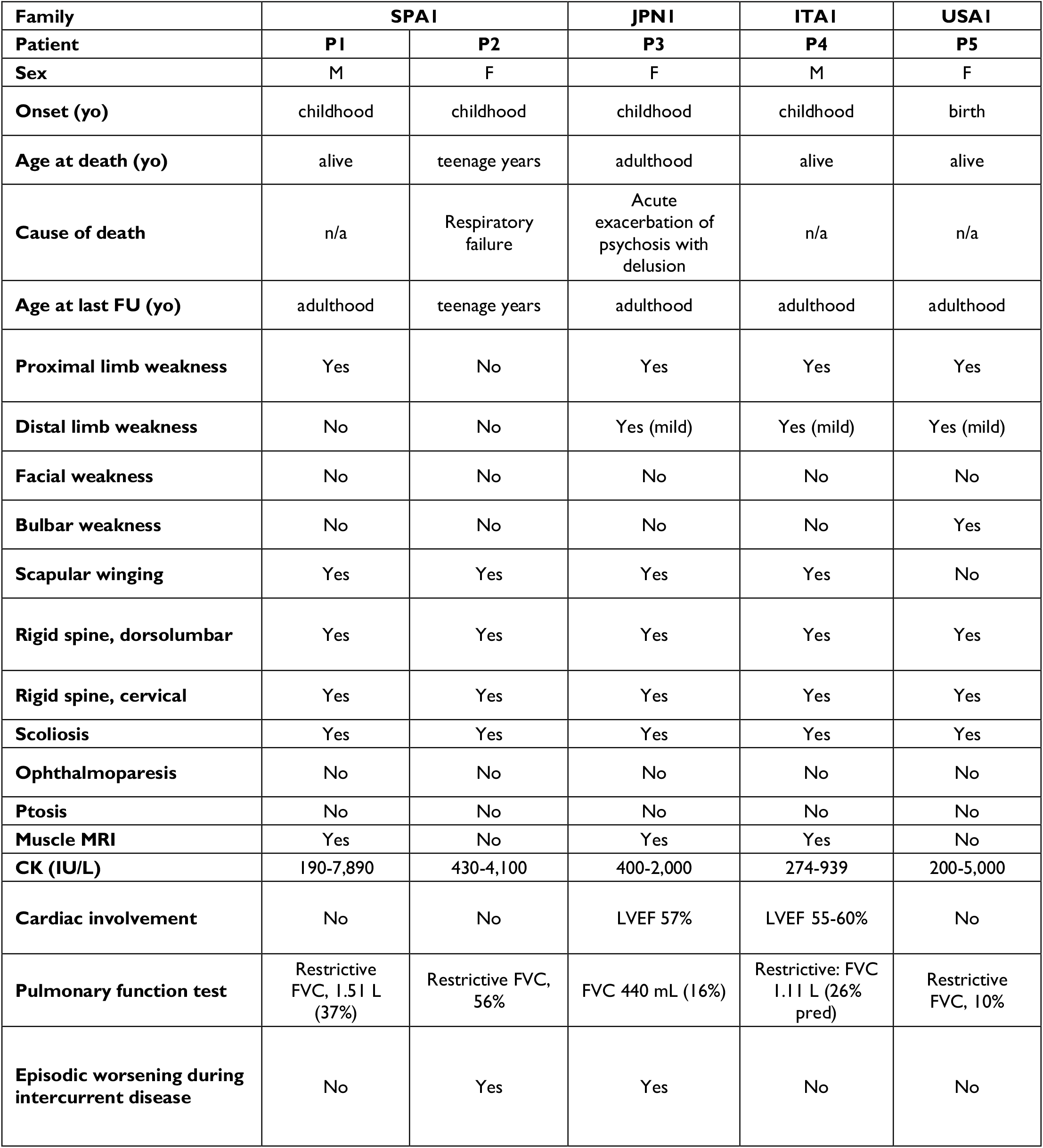
Clinical summary of the HMGCS1-myopathy cohort.

Muscle MRI performed for two of five patients (P1, P4) showed variability in T1 signaling primarily in the posterior muscles of the hip, thigh, and leg (Fig 2A). Muscle MRI of patient P1 (SPA1) indicated extensive fatty replacement in the cervical paraspinal muscles (Fig 2Ai) as well as involvement of the posterior compartment of the thighs (Fig 2Aii). Edema was also observed in the posterior compartment of the thighs on STIR (Short Tau Inversion Recovery) imaging of patient P1 (Fig 2Aiii). Lower leg muscle MRI of patient P1 was normal (not shown). Muscle MRI of patient P4 (ITA1) similarly indicated fatty infiltration in the posterior compartment of the thighs (Fig 2Aiv,v) as well as in the posterior compartment of the leg (Fig 2Avi).

**Figure 2.**
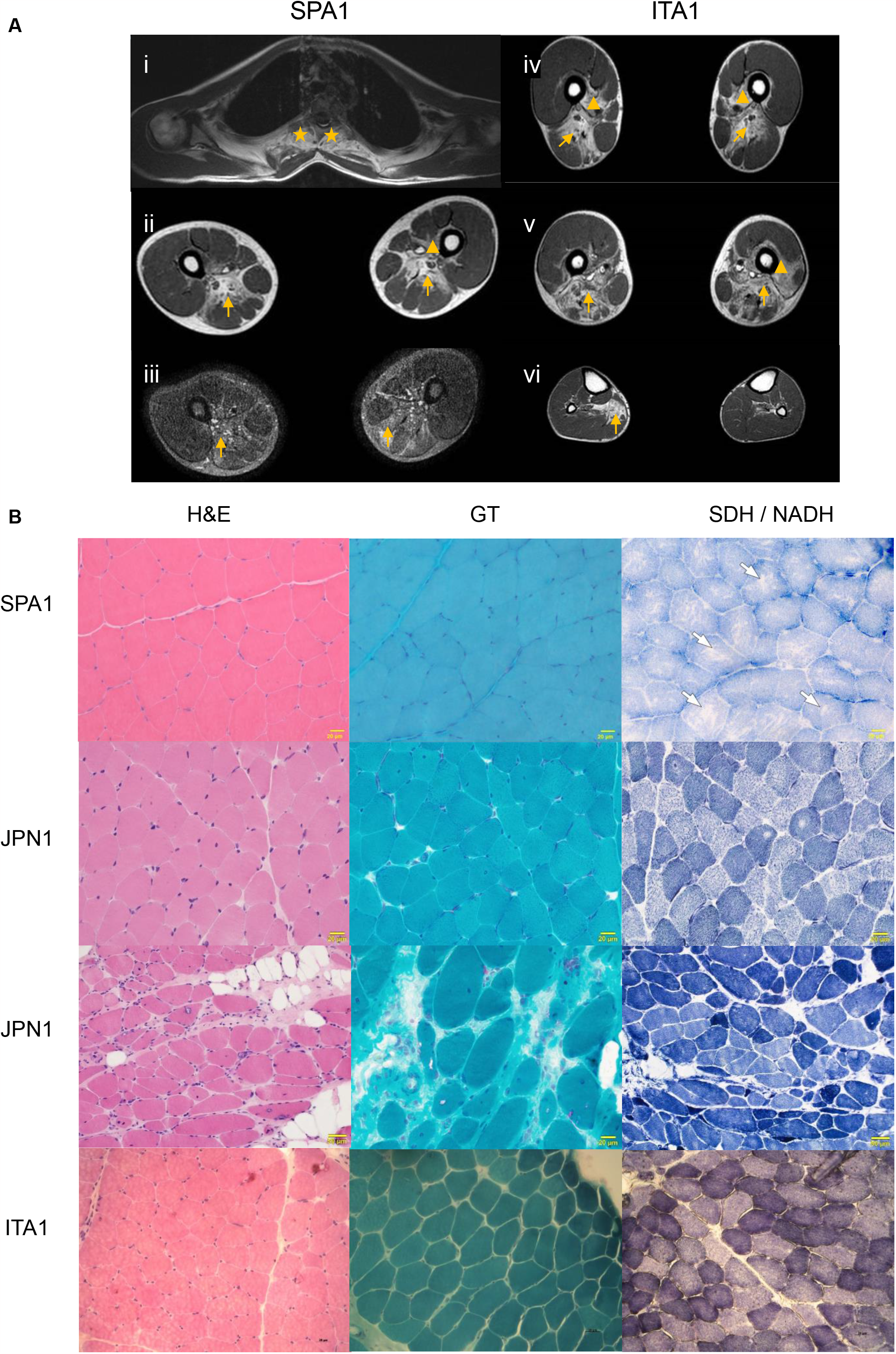
Muscle MRI and pathology associated with HMGCS1-myopathy. **(A)** Muscle MRI performed for Patient P1 (SPA1) (i-iii) and Patient P4 (ITA1) during adulthood (iv-vi). Variation in T1 signal indicative of fatty replacement in the posterior muscles of the cervical paraspinal muscles (stars) (i), the posterior compartment of the thigh (arrows) (ii, iv, v), and the posterior compartment of the leg (arrows) (vi). Evidence of edema in the posterior compartment of the thighs in Patient P1 shown by STIR (arrows) (iii). Vastus intermedious is also affected to a lesser extent (arrow heads). **(B)** Light microscopy staining of muscle biopsies from patients P1 (SPA1) during teenagerhood (top panel), P3 (JPN1) during childhood and adulthood (middle panels), and P4 (ITA1) during childhood (bottom panel). Haematoxylin & Eosin (H&E) and Gömöri Trichrome (GT) staining show occasional internal nuclei and rimmed vacuoles in some patients. Succinate dehydrogenase (SDH) staining of SPA1 and nicotinamide adenine dinucleotide (NADH) staining of JPN1 and ITA1 patient biopsies reveal irregularities in staining including moth-eaten appearances and core-like regions (arrows). Scale bars represent 20 μm or 50 μm as indicated.

Four patients (P1, P3, P4, P5) underwent muscle biopsy. The most noticeable findings were irregularities on oxidative enzyme staining, including core-like regions in some myofibres as shown by SDH and NADH staining (Fig 2B). There were also occasional internal nuclei and rimmed vacuoles observed on H&E and Gömöri Trichrome staining (Fig 2B) in some patients. The muscle pathology of patient P3 worsened with age, showing the presence of rimmed vacuoles and dystrophic features during adulthood which were absent during childhood (Fig 2B). Muscle biopsy for patient P5 was indicative of a chronic myopathy with marked macrophage infiltration. There were also vacuoles and atrophic myofibres present (images unavailable).

### Genetic investigations

We identified seven novel or rare biallelic variants in *HMGCS1* (ENSG00000112972) co-segregating with rigid spine syndrome in four families (Fig 1A, Fig 3A). This included six missense variants and one frameshift variant. In the index family (SPA1), we identified a homozygous missense variant in *HMGCS1*, c.1339T>C, p.(Ser447Pro) in the two affected siblings (P1, P2). The variant was absent in gnomAD data (v2.1.1) and was heterozygous in the unaffected parents as confirmed by Sanger sequencing. Additional rigid spine syndrome cases with biallelic *HMGCS1* variants were subsequently identified via international collaborations and gene-matching platforms, including the Broad Institute’s matchbox. In JPN1, exome data for patient P3 revealed a maternally inherited missense variant (c.86A>T, p.(Gln29Leu)) in trans with a paternally inherited frameshift variant (c.344_345del, p.(S115Wfs*12)). The two unaffected siblings were identified as carriers for the frameshift variant (Fig 1A). In ITA1, the affected proband (P4) was identified with compound heterozygous missense variants (c.890G>T, p.(Gly297Val) maternal allele, in trans with c.1289G>A, p.(Arg430Lys), paternal allele). In USA1, patient P5 had compound heterozygous missense variants: c.209T>C, p.(Met70Thr), maternal allele, in trans with c.803G>C, p.(Cys268Ser), paternal allele. All variants identified in this cohort were absent from gnomAD except for the c.803G>C, p.(Cys268Ser) variant, which was present in the heterozygous state in 4 of 251,086 alleles (all from the European non-Finnish population, frequency 1.59x10^−5^). Apart from the M70 residue, all substituted residues in this cohort are conserved in multiple species as well as in human HMGCS2, the mitochondrial paralogue (Fig 3B). Further details including variant effect predictions are included in Supplementary Table 1.

**Figure 3.**
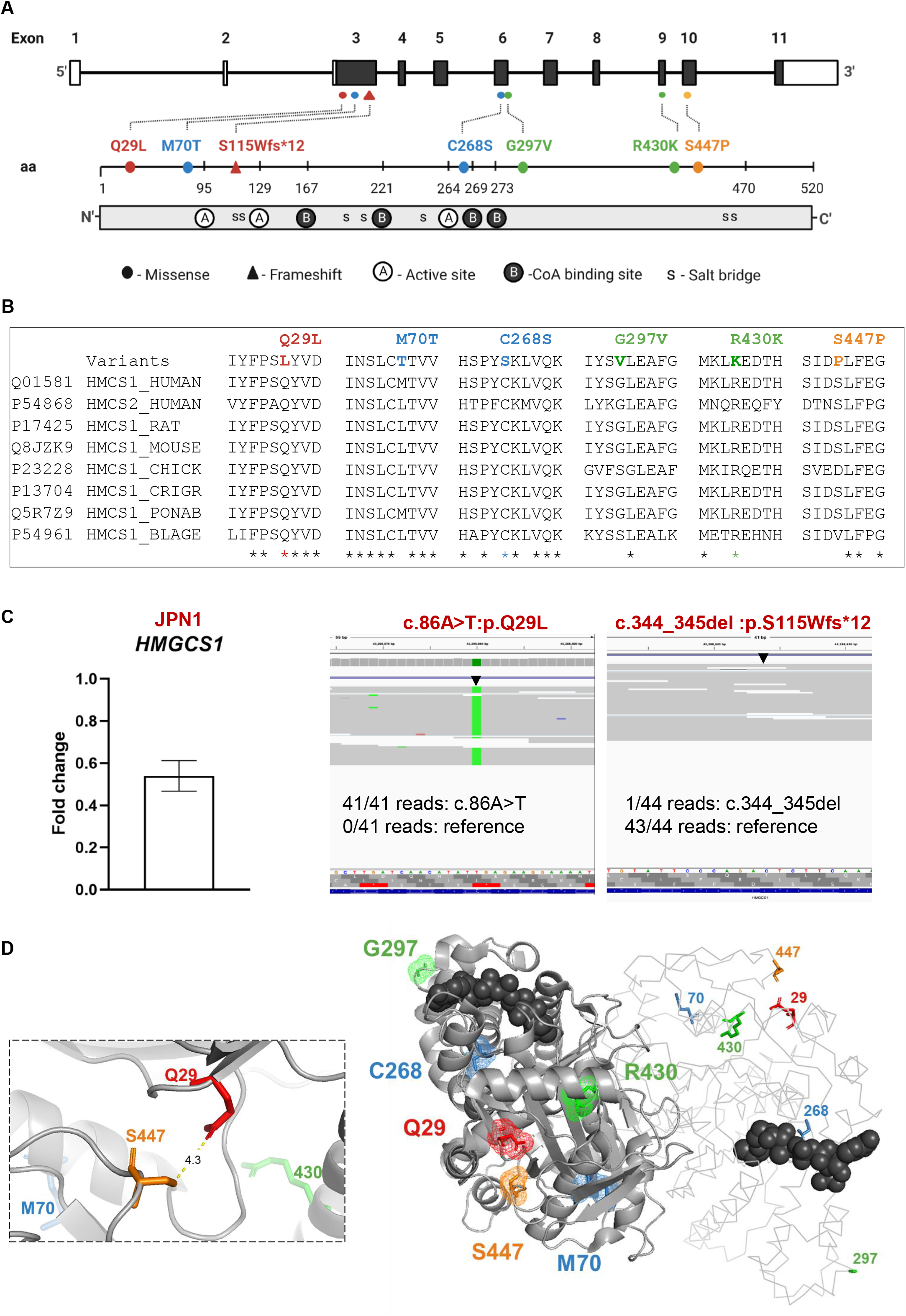
*HMGCS1* variants. **(A)** Schematic representations of the *HMGCS1* gene (top) and protein (bottom) linear architectures. The *HMGCS1* gene includes untranslated regions (empty boxes), coding exons (filled boxes) and introns (horizontal line). Variants identified in patients with rigid spine syndrome are labelled, including both missense (circle) and truncating (triangle) variants. The variants are coloured according to the families they have been identified in (red: JPN1, blue: USA1, green: ITA1, orange: SPA1). Letters A, B and s within the protein structure (bottom) respectively represent residues of the active site (E95, C129, H264), coenzyme A binding site (N167, S221, K269, K273) and salt bridge (D119, E121, R194, D208, K239, K461 and H462). Diagram not to scale. **(B)** Conservation of the six missense variant positions in various species and in HMGCS2; the mitochondrial paralogue. Asterisks indicate complete conservation of residues in the analysed species. **(C)** RNA-seq fold change in *HMGCS1* expression in patient P3 (JPN1) compared to three unaffected controls (left). Error bars correspond to standard deviation. RNA-seq reads indicate virtually monoallelic expression of *HMGCS1* c.86A>T, p.(Q29L) suggesting the c.344_345del, p.(S115W*fs*\*12) variant undergoes nonsense mediated decay (right figure). **(D)** Visualisation of the HMGCS1 dimer (PDB: 2P8U) using PyMOL. HMGCS1 chain A (right) represented by a ribbon and chain B (left) shown as a cartoon. Ac-CoA represented as dark grey spheres. Variant positions (represented by licorice sticks) coloured by family. Close-up representation showing the minimum distance between S447 and Q29 is ∼4.3 Å (dashed box).

The c.209T>C, p.(Met70Thr) variant was also identified via gene matching in an additional proband (in the USA, Supplementary Table 1, USA2) presenting with a similar unresolved rigid spine phenotype. This variant was heterozygous in the proband and inherited from the unaffected father. However, whole genome sequencing and RNA-seq of patient fibroblasts did not identify a second likely pathogenic *HMGCS1* variant in trans for this patient suggesting a possible more complex underlying genetic cause such as a genomic rearrangement. Alternative, it remains inconclusive whether *HMGCS1* is the underlying genetic cause of rigid spine syndrome for this case.

The compound heterozygous frameshift variant (c.344_345del, p.(Ser115Trpfs*12)) in patient P3 (JPN1) was predicted to result in nonsense mediated decay of the transcript. Indeed, RNA-seq of patient muscle showed only one read (out of 44) contained the frameshift variant while all reads (41/41) spanning the c.86 position contained the c.86A>T, p.(Gln29Leu) variant (Fig 3C). In further support, expression analyses via DEseq^29^ indicated there was a 0.53 fold decrease in *HMGCS1* transcript levels in the patient relative to three controls (Fig 3C). Collectively, this suggests that at a protein level, in patient P3, the c.86A>T, p.(Gln29Leu) variant is almost exclusively present.

To determine whether HMGCS1 dysfunction is associated with aberrant expression of other genes of the mevalonate pathway, we analysed RNA-seq for patient P3 against 129 individuals by OUTRIDER.^32^ We identified eight expression outliers in patient P3 (Supplementary Fig 1). However, no candidates showed any remarkable associations with the mevalonate pathway or other shared pathway according to gene ontology enrichment analyses.

### Protein studies

The variants identified in the patients were scattered in several exons in *HMGCS1* (Fig 3A). These include exon 3 (p.(Q29L), p.(M70T), p.(S115Wfs*12)), exon 6 (p.(C268S), p.(G297V)), exon 9 (p.(R430K)) and exon 10 (p.(S447P)). The C268S substitution is adjacent to a coenzyme A (CoA) binding residue, K269 (Fig 3A). Positions of the remaining substitutions were distant from residues of the CoA active sites (E95, C129, H264), binding sites (S221, K269, K273) and residues of the dimer salt bridge (D119, Glu121, R194, D208, K239, K461 and H462). Three substituted residues (Q29, M70, and S447) appear to cluster towards the surface of HMGCS1 (Fig 3D). Residues Q29 and S447 appear <5 Å apart in 3D space according to the PDB model (Fig 3D).

### *HMGCS1* is abundant in various tissues, including developing and adult skeletal muscle

RNA-seq data from the NCBI BioProject (PRJEB4337)^48^ and the GTEx portal^49^ indicated that *HMGCS1* is enriched in brain and liver and is expressed at lower levels in other tissues, including skeletal muscle. Similarly, qPCR analyses indicated enriched expression of *HMGCS1* in human cortex, and relatively lower expression in cells and tissues, including myotubes and adult skeletal muscle (Fig 4A). Of the myogenic samples analysed in the FANTOM5^28^ database, *HMGCS1* expression was highest in skeletal muscle satellite cells (235-462 transcripts per million (TPM)) relative to fetal-derived skeletal muscle cells (3-326 TPM), myotubes (0-240 TPM), and myoblasts (31-50 TPM).

**Figure 4.**
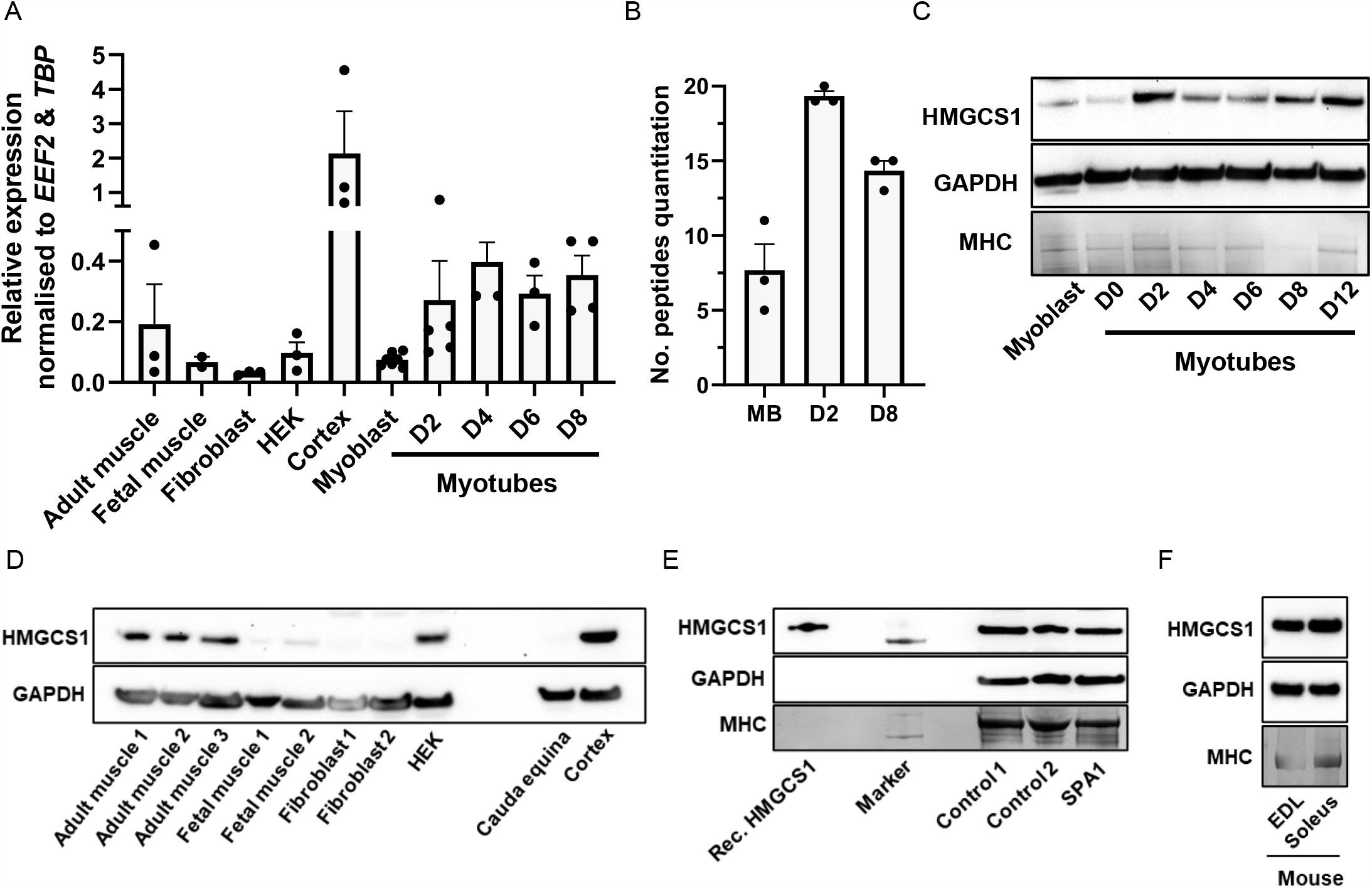
*HMGCS1* is expressed in various tissues including patient skeletal muscle. **(A)** Quantitative real-time PCR analysis using cDNA from healthy human cell lines and tissues. Expression data from human embryonic kidney cells (HEK293FT cell line), fibroblasts, myoblasts (D0), myotubes at D2, D4, D6, D8 of differentiation, cortex, skeletal muscle controls from *in vitro* contractile testing and fetal muscle. Transcript levels were normalised to the geometric mean of *EEF2* and *TBP* using the delta Ct method. Data presented as mean ± SEM (*n* =2-7 biological). **(B)** Graphical presentation of HMGCS1 peptides detected by quantitative mass spectrometry from myoblasts (MB) and myotubes at day 2 (D2) and day 8 (D8) of differentiation. **(C-F)** Western blotting for HMGCS1 (top panels) in **(C)** primary human (Cook Myosite) myoblasts and myotubes from days 0-12 of differentiation, in **(D)** control human tissue and cell lines, and in **(E)** patient P1 (SPA1) skeletal muscle alongside healthy skeletal muscle controls. Recombinant wildtype HMGCS1 (Rec. HMGCS1) used as a control for antibody specificity. **(F)** Western botting for HMGCS1 in mouse *extensor digitorum longus* (EDL) and soleus muscles (top panel). Western blots for GAPDH and gel-stained myosin heavy chain (MHC) bands are shown to demonstrate comparable loading of total and muscle proteins.

Western blotting for HMGCS1 in human tissues and cells revealed a band of the expected size (∼57 kDa; Fig 4C-F), which matched the product size of the recombinant HMGCS1 protein (Fig 4E). We show that HMGCS1 is enriched in the cortex (Fig 4D), consistent with the qPCR data and the proteomic data from the Human Protein Atlas^50^. In addition, HMGCS1 was detected in adult human skeletal muscle and was relatively less abundant in human fetal skeletal muscle (Fig 4D), consistent with the qPCR studies. HMGCS1 was also detected in human myoblasts and myotubes from days 0 to 12 of differentiation by mass spectrometry (Fig 4B) and/or western blotting (Fig 4C). There was comparable abundance of HMGCS1 in skeletal muscle of patient P1 (SPA1) and control skeletal muscle biopsies (Fig 4E) suggesting the p.(S447P) substitution does not reduce protein stability or abundance. Similar levels of Hmgcs1 were detected in mouse EDL (*extensor digitorum longus*) and soleus muscles (Fig 4F), suggesting that Hmgcs1 is similarly abundant in fast- and slow-twitch myofibres in mice.

### Functional investigations of recombinant HMGCS1

HMGCS1 has been reported to exist as a dimer.^25,51^ To investigate whether substitutions in HMGCS1 affect dimerisation, the recombinant wildtype (HMGCS1^WT^) and mutant proteins (HMGCS1^S447P^, HMGCS1^Q29L^, HMGCS1^M70T^ and HMGCS1^C268S^) were analysed by size exclusion chromatography to evaluate monomer and dimer content. All proteins eluted at equivalent points corresponding to the expected molecular weight for dimerised HMGCS1 (Supplementary Fig 2A,B). This suggested that the substitutions do not interfere with HMGCS1 dimerisation. There were second peaks detected for HMGCS1^WT^ and HMGCS1^Q29L^ (Supplementary Fig 2A), however, their elution volumes did not correspond to the expected molecular weight of the monomer (Supplementary Fig 2B).

Circular dichroism spectra and melting curve analyses were performed to investigate whether the HMGCS1 variants affected protein structure and thermal stability, respectively. Overall, the wildtype and all mutants investigated appeared structurally stable, showing considerable overlap in the CD spectra (190−260 nm) throughout the melting range (15−95°C; Fig 5A-D). The majority of the protein remained intact even at high temperatures (> 60°C), showing marginal changes in CD signal suggesting the proteins have a thermally stable core (Fig 5E). The melting curves we measured suggested only a small part of the protein unfolded below 95°C. Owing to the relative stability of the proteins, the small changes in CD signal, and the variability between experiments, there were no statistically significant differences in protein stability detected between the mutants and the wildtype (one-way ANOVA, Fig 5F). Though there was a trend suggesting the M70T and C268S mutants may be less stable than the wildtype across the independent experiments (Fig 5F). The WT and p.S447P proteins were run in three additional experiments in which the p.S447P protein showed a small (2.6°C) decrease in protein stability compared to WT (paired t-test, *n* = 7, *p* = 0.03; Fig 5F).

**Figure 5.**
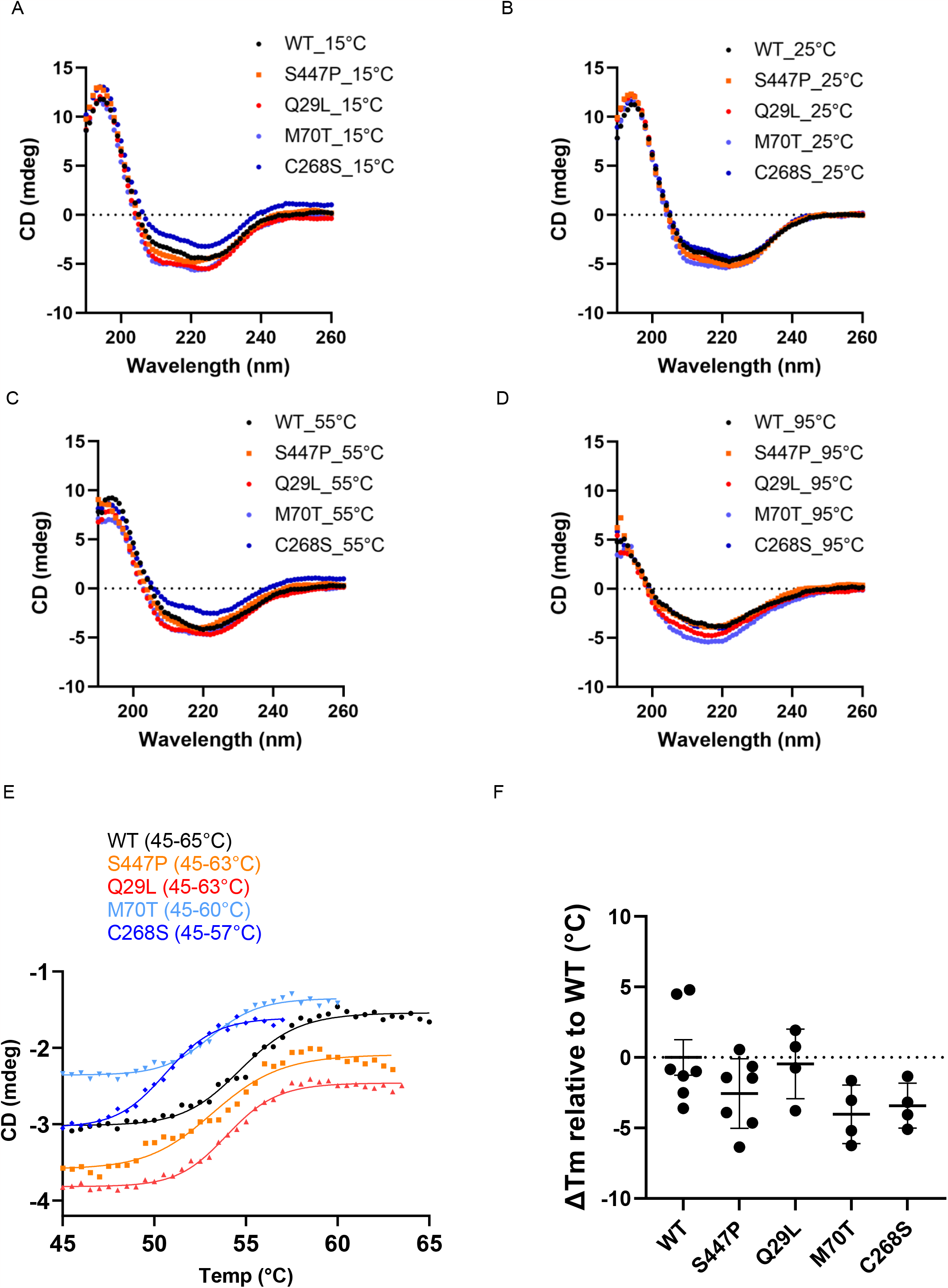
Circular dichroism analyses of recombinant wildtype and mutant HMGCS1. **(A-D)** Circular dichroic spectra scans (195-260 nm) of wildtype and mutant HMGCS1 (0.1 mg/mL) measured at **(A)** 15°C, **(B)** 25°C, **(C)** 55°C and **(D)** 95°C in buffer containing 10 mM sodium phosphate buffer pH 7.4, (including 1% of original protein buffer: 150 mM NaCl, 50 mM Tris and 10% glycerol). **(E)** Representative melting curves of wildtype and mutant HMGCS1 (0.1 mg/mL) measured at 229 nm. Melting curves (37-55°C) were fitted to sigmoidal logistic regression four parameter fits where the IC50 (point of inflection) were taken as the melting temperatures (T_m_) plotted in **(F)**, data (*n*=4-7) presented as mean ± SEM.

All mutants also appeared to retain enzymatic activity and showed no significant changes in activity compared to the wildtype when assayed under multiple substrate concentrations (Supplementary Fig 3). This included HMGCS1^C268S^ which, despite harboring a substitution neighboring the CoA binding site, only showed a ∼30% reduction in activity relative to the wildtype when assayed with 200 μM AcAc-CoA and Ac-CoA (Supplementary Fig 3B). However, the small changes in the absorbance and the resulting variability between runs made it difficult to confirm whether any partial changes detected were significant. Nonetheless it was apparent that the substitutions do not drastically impair enzymatic activity.

### *hmgcs1* null zebrafish present severe movement defects, mitochondrial abnormalities, and an early lethal phenotype

To evaluate the effect of loss of HMGCS1 function in an animal model during developmental stages we generated a *hmgcs1* mutant zebrafish, containing a 302 bp deletion in exon 3, which results in a frameshift after 35 aa (Supplementary Figure 5A). Loss of Hmgcs1 in zebrafish embryos results in severe early developmental defects, with *hmgcs1*^*-/-*^ embryos displaying blood pooling in the brain, reduced pigmentation (Figure 6A), absence of movement at 48 hours post fertilisation (hpf, Fig 6B), and lethality at 3 days post fertilisation (dpf, Fig 6C). Further investigation of the movement defect identified that *hmgcs1*^*-/-*^ embryos completely lack a touch-evoked escape response^52^ compared to *hmgcs1*^*+/+*^ and *hmgcs1*^*+/-*^ siblings.

**Figure 6.**
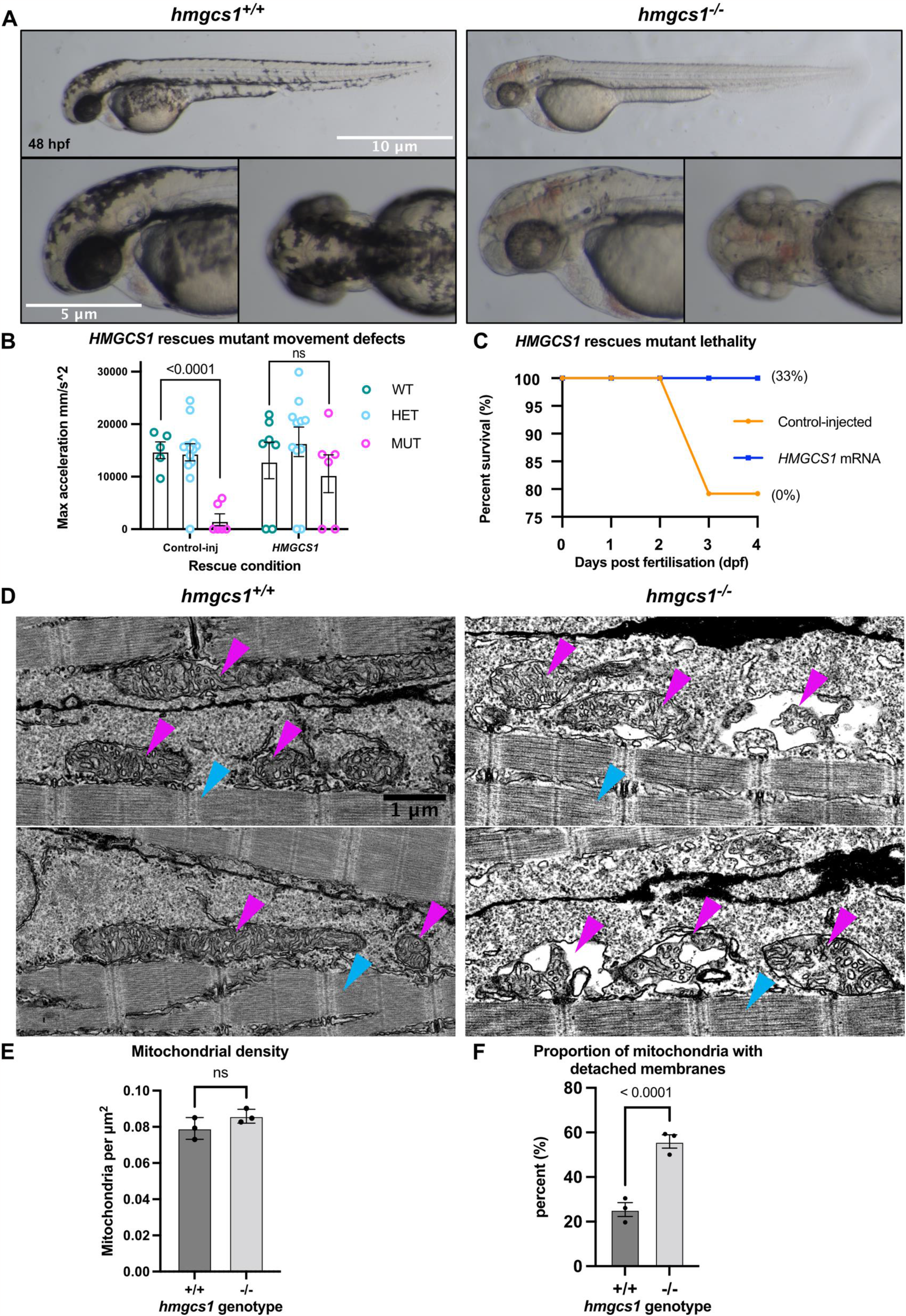
*hmgcs1*^*-/-*^ zebrafish have severe mitochondrial abnormalities and an early lethal phenotype. **(A)** Brightfield images of wildtype and *hmgcs1*^*-/-*^ embryos at 48 hours post fertilisation (hpf). *hmgcs1*^*-/-*^ embryos have developmental delay, reduced pigmentation, blood pooling in the brain. **(B)** Quantification of movement to a touch-evoke assay. *hmgcs1*^*-/-*^ embryos lack a movement response to touch, unlike wildtype and heterozygous siblings. Injection of mRNA encoding HMGCS1 rescues the movement defects in *hmgcs1*^*-/-*^ embryos. **(C)** Survival assay on the progeny of a *hmgcs1*^*+/-*^ in cross. In control injected fish only 79% survived until day 4 and no *hmgcs1*^*-/-*^ fish were detected at this time, as indicated in brackets, demonstrating that loss of Hmgcs1 results in early embryo lethality. Injection of *HMGCS1* mRNA rescues the lethality in *hmgcs1*^*-/-*^ embryos, with 100% of injected fish surviving until day 4, and subsequent genotyping indicating 33% of the fish were *hmgcs1*^*-/-*^. 24 embryos from the *hmgcs1*^*+/-*^ in cross were used for each assay. **(D)** Scanning electron microscopy images of longitudinal muscle sections at 48hpf, demonstrating mitochondria associated with skeletal muscle. *hmgcs1*^*-/-*^ embryos have an increased proportion of mitochondria with detached membranes. Magenta arrows indicate mitochondria, cyan arrows indicate myofibres. Scale bar represents 1 μm. **(E)** Mitochondrial density is not affected in *hmgcs1*^*-/-*^ embryo skeletal muscle (students t-test). **(F)** The proportion of mitochondria with detached membranes is higher in *hmgcs1*^*-/-*^ embryos compared to wildtype (Fisher’s exact test). Statistics were performed in Prism 9 (GraphPad). Statistically significant differences are indicated on the graphs by the p-value.

Injection of mRNA encoding HMGCS1 was able to rescue early lethality in *hmgcs1*^*-/-*^ embryos (Figure 6C), as well as the movement defects in response to touch (Figure 6B), demonstrating that these defects specifically result from the loss of Hmgcs1 function.

Electron microscopy to investigate the muscle ultra structure revealed that *hmgcs1*^*-/-*^ embryos exhibited significant mitochondrial abnormalities within the skeletal muscle at 48 hpf, presenting as a detachment or invagination of the inner membrane (Figure 6D,F), without affecting mitochondrial density (Figure 6E). Interestingly, the myofibril pattern in *hmgcs1*^*-/-*^ animals was indistinguishable from wildtype siblings, indicating that the muscle structure was not affected (Figure 6D, cyan arrows). These results indicate that absence of Hmgcs1 results in damage to the integrity of the mitochondrial membrane and loss of muscle function.

## Discussion

We report the identification of seven biallelic variants in *HMGCS1* in four unrelated families with rigid spine syndrome. Although the cohort thus far includes five patients, the distinct rigid spine phenotype shared by the patients, including disease onset and severity suggests a common genetic basis of disease which we define here as *HMGCS1*-related myopathy.

*HMGCS1* encodes the cytosolic HMG-CoA synthase; a key enzyme of the mevalonate pathway which catalyses the condensation reaction between AcAc-CoA and Ac-CoA to form HMG-CoA.^25^ Its mitochondrial paralogue; HMGCS2 catalyses the same reaction for the ketogenesis pathway. Biallelic variants in *HMGCS2* are associated with HMG-CoA synthase deficiency (OMIM# 605911) characterised by episodic metabolic distress, febrile crisis, hepatomegaly, and respiratory distress.^53-55^ HMGCS1 and HMGCS2 share ∼67% sequence similarity and five of the six substituted HMGCS1 residues presented here (Q29, C268, G297, R430 and S447) are conserved in HMGCS2 (Q66, C305, G334, R467 and S484, respectively) suggesting that they are of functional importance. Thus far, none of the HMGCS1 substitutions observed in this study overlap with the reported HMGCS2 pathogenic substitutions. Nevertheless, the association of biallelic variants in *HMGCS2* with disease suggests that biallelic variants in *HMGCS1* can be similarly deleterious. This is further supported by gnomAD constraint scores which indicate that *HMGCS1* (pLI=1; Z=3.04) is substantially more intolerant to loss-of-function and missense variation compared to *HMGCS2* (pLI=0; Z=-0.19).

Myopathic and rigid spine phenotypes have been associated with several inborn and acquired errors of the mevalonate pathway.^11,20,21,56-64^ This includes biallelic variants in *HMGCR* (HMG-CoA reductase) immediately downstream of HMGCS1 in the pathway (Supplementary Fig 4), which have recently been associated with limb-girdle muscular dystrophy (OMIM# 620375).^21,65^ Inhibition of HMGCR by statins and autoantibodies are also associated with acquired muscle disorders including statin-induced myopathy^20^ and anti-HMGCR myopathy.^59,61,66,67^ Further downstream, inhibition of FDPS (farnesyl diphosphate synthase) by bisphosphonate medications has been associated with profound muscle weakness with fever and flu-like symptoms.^68,69^ Moreover, biallelic variants in *GGPS1* have been associated with a muscular dystrophy (OMIM# 619518).^11,70^ Of note, *GGPS1*-related muscular dystrophy includes several overlapping phenotypes including scoliosis, joint contractures, hypotonia, elevated creatine kinase, as well as episodic worsening of muscle weakness and respiratory insufficiency during intercurrent disease.^11,70^ Overall, the myopathic phenotypes implicated in the mevalonate pathway support *HMGCS1* as an underlying genetic cause for our patients’ disorders.^11^

The mevalonate pathway branches into several critical other pathways including the cholesterol biosynthesis pathway, the isoprenoid pathway and the ubiquinone pathway, which are essential for regulating cell proliferation, maturation, and maintenance.^71^ Genes of these pathways, including *HMGCS1* are conserved in eukaryotes, archaea and some bacteria.^25,71-73^ Knockdown of most genes of the mevalonate pathway is associated with complete preweaning lethality in mice (Supplementary Fig 4).^11,74^ Knockdown of *Hmgcs1* in mice is associated with prenatal lethality (International Mouse Phenotyping Consortium, https://www.mousephenotype.org/data/genes/MGI:107592), with no homozygous pups observed at E9.5. It is thought that this arises due to placental defects (personal communication, JAX lab). A severe phenotype was similarly observed in our studies of *hmgcs1*^*-/-*^ zebrafish, which included developmental delay, immobility by 2 dpf, and fatality during 3 dpf. There were also significant mitochondrial abnormalities in the skeletal muscle, which is consistent with mitochondrial changes reported in a *c. elegans hmgr-1* mutant.^75^ Pharmacological inhibition of Hmgcr in zebrafish and mouse models is associated with severe multi-system defects despite predominantly being associated with myopathies in humans.^76-78^

Given the severity and developmental onset of disease in animal models resulting from loss of HMGCS1, it is perhaps not surprising that the variants identified in patients do not dramatically impair HMGCS1 function. We suspect that biallelic loss-of-function variants in humans would cause prenatal lethality consistent with the anomalies observed in animal studies.^79^ There are no individuals in gnomAD homozygous for loss-of-function HMGCS1 variants and only four individuals with heterozygous loss of function variants, supporting that HMGCS1 function is critical to life.

We have functionally investigated four of six HMGCS1 substitutions (S447P, Q29L, M70T and C268S) identified in our rigid spine syndrome cohort and showed no dramatic functional differences between the mutants and the wildtype. All mutants retained their 3D structure and dimerisation tendencies according to CD spectroscopy and size exclusion chromatography. These findings were similar to other studies of recombinant HMGCS1 mutants (p.C129A; *H. sapiens*^80^ and p.A110G; *E. faecalis*^*51*^) showing no major difference in HMGCS1 structure or conformation. However, subtle albeit significant changes were detected by X-ray crystallography.^51^ We thus speculate that the structural effect of the substitutions observed in our cohort (if any) may be partial and local, and therefore beyond the detection capacity of CD. Interestingly, comparison with an AcAc-CoA thiolase/HMGCS complex structure from *Methanothermococcus thermolithotrophicus* reveals that the residues equivalent to the Q29L, M70T and S447P substitutions are located on a region that interacts with a scaffold protein (Pfam DUF35).^81^ Disruption of such a complex in humans could further explain the effect of these mutations, however, no scaffold protein and ternary complex has yet been described for human AcAc-CoA thiolase/HMGCS.

We initially predicted that the HMGCS1 substitutions may impair HMGCS1 activity, given that HMGCS2 substitutions, including those distant from the catalytic site, have been associated with reduced enzymatic activity.^79^ All four HMGCS1 mutants maintained enzymatic function within the same order of magnitude as the wildtype, including HMGCS1^C268S^ which harbours a substitution close to the active site. This mutant maintained ∼70% of the activity of the wildtype. Of note, a superficial substitution in HMGCS2 p.R505Q (p.R468 in HMGCS1) has been associated with disease despite similarly retaining 70% of normal HMGCS2 activity.^79^ Pathogenic substitutions in *GGPS1* have also been reported to have no significant impact on GGPS1 conformation and only mildly impaired enzyme activity *in vitro* and in patient-derived fibroblasts.^11^ Similarly we see that hypomorphic variants in *HMGCS1* show activity levels similar to WT using *in vitro* assays. Some suggest that subtle changes in enzymatic function can lead to abnormal accumulation and processing of metabolites, causing dysregulated activities of other enzymes of the mevalonate pathway.^56,72,82^

We have demonstrated that *HMGCS1* is expressed and detected in myoblasts, myotubes and skeletal muscle. Although enrichment of *HMGCS1* in the brain may suggest a neurological phenotype should result. *HMGCR* and *GGPS1* are also more abundant in brain tissue compared to skeletal muscle^28,48-50^ but have been predominantly associated with myopathic phenotypes.^11,59,60,83-87^ By western blotting, Hmgcs1 was similarly detected in mouse EDL and soleus muscle, suggesting that Hmgcs1 is similarly abundant in fast- and slow-twitch myofibres.

There are various hypotheses that suggest how disruptions to the mevalonate pathway cause skeletal muscle disease.^20,88^ These include alterations in the cholesterol abundance and fluidity of the sarcolemma, impaired mitochondrial function, and dysregulated isopentylation of selenocysteine-tRNA, which is required for *SELENON* expression.^20,89,90^ Of note, the clinical resemblance between patients with *SELENON*-related and *HMGCS1*-related rigid spine syndrome, as well as other myopathies associated with the mevalonate pathway, may suggest dysregulated isopentylation of selenocysteine-tRNA as a strong candidate pathomechanism.^10^ An alternative explanation to the muscle involvement may be associated with a dysregulation of isoenzymes of the mevalonate pathway exclusively expressed in skeletal muscle.^91,92^ For example, IDI2 (isopentenyl diphosphate isomerase 2; Supplementary Fig 4) is a skeletal muscle-specific isozyme of IDI1 which appears to be regulated by HMGCR abundance and activity.^92^ Statin inhibition of Hmgcr has been associated with significantly reduced *Idi2* mRNA levels in mouse skeletal muscle, whereas *Idi1* expression remained unaffected.^92^ This suggests that dysregulation of muscle-specific isoenzymes may differentially affect the mevalonate pathway in skeletal muscle compared to other tissue.^92^ Disruption of the mevalonate pathway, whether through statin inhibition^93^ or mutation^75^, also leads to mitochondrial abnormalities, consistent with the findings observed in *hmgcs1*^*-/-*^ zebrafish mutants. Interestingly, administration of mevalonic acid (the metabolite downstream of hmgcr) has been effective in treating HMGCR-myopathy in humans^75^ and statin induced myopathy in mice.^76,94,95^ Overall, the disease mechanisms underlying the muscle phenotype may be clarified by multi-omic analyses as additional patients with mevalonate pathway-related myopathies are identified.^96^

## Conclusion

We report a novel autosomal recessive rigid spine syndrome we have termed *HMGCS1*-myopathy. We speculate that *HMGCS1* variants act by a hypomorphic mechanism, however, the precise cause of the myopathic phenotype in the patients remains to be elucidated. This report expands the genetic causes of rigid spine syndrome and highlights the mevalonate pathway as an essential pathway for muscle function. We encourage screening of *HMGCS1* in patients with molecularly unresolved rigid spine syndrome.

## Data Availability

The data and constructs generated by this study are available upon request.

## Abbreviations

AcAc-CoA: acetoacetyl coenzyme A
Ac-CoA: acetyl coenzyme A
CD: circular dichroism
HMG-CoA: 3-hydroxy-3-methylglutaryl coenzyme A
K_m_: Michaelis constant
MRI: magnetic resonance imaging
TPM: transcripts per million
V_max_: maximum velocity
WES: whole exome sequencing
WGS: whole genome sequencing

## Acknowledgements

We would like to thank the patients and families for their participation in this study. We thank Dr. Amelie Nadeau and Dr. Katy Meilleur for their clinical expertise.

## Supplementary material

Supplementary material is available online.

