## Supplementary material for "Biallelic variants in *HMGCS1* are a novel cause of rare rigid spine syndrome"

### Clinical summaries

Clinical summaries for each case are available upon request.

**Supplementary Table 1 Summary of the biallelic variants in *HMGCS1* identified in the HMGCS1-myopathy cohort**

| Family | Consanguineous | Variant (NM_001098272.2) | ClinVar Accession # | gnomAD | CADD | PolyPhen-2 | Provean | SIFT |
|---|---|---|---|---|---|---|---|---|
| SPA1 | N* | A, B. c.1339T>C, p.(Ser447Pro)[Hom] | SCV004036160 | Absent | 23.3 | Benign | Neutral | Tolerated |
| JPN1 | N | A. c.86A>T, p.(Gln29Leu) | SCV004036161 | A. Absent | A. 23.1 | A. Benign | A. Deleterious | A. Tolerated |
| | | B. c.344_345del, p.(Ser115Trpfs*12) | SCV004036162 | B. Absent | | | | |
| ITA1 | N | A. c.1289G>A (p.Arg430Lys) | SCV004036163 | A. Absent | A. 26.1 | A. Possibly damaging | A. Deleterious | A. Damaging |
| | | B. c.890G>T (p.Gly297Val) | SCV004036164 | B. Absent | B. 23.7 | B. Possibly damaging | B. Deleterious | B. Tolerated |
| USA1 | N | A. c.803G>C, p.(Cys268Ser) | SCV004036165 | A. $1.59 \times 10^{-5}$ | A. 27.7 | A. probably damaging | A. Deleterious | A. Tolerated |
| | | B. c.209T>C, p.(Met70Thr) | SCV004036166 | B. Absent | B. 23.9 | B. Benign | B. Deleterious | B. Damaging |
| USA2 | N | A. c.209T>C, p.(Met70Thr) | SCV004036166 | A. Absent | A. 23.9 | A. Benign | A. Deleterious | A. Damaging |
| | | B. no second variant detected | | | | | | |

[A]First allele.
[B]Second allele.
*Parents were from a small village. Consanguinity suspected but denied.
