## Supplementary figures for "Biallelic variants in *HMGCS1* are a novel cause of rare rigid spine syndrome"

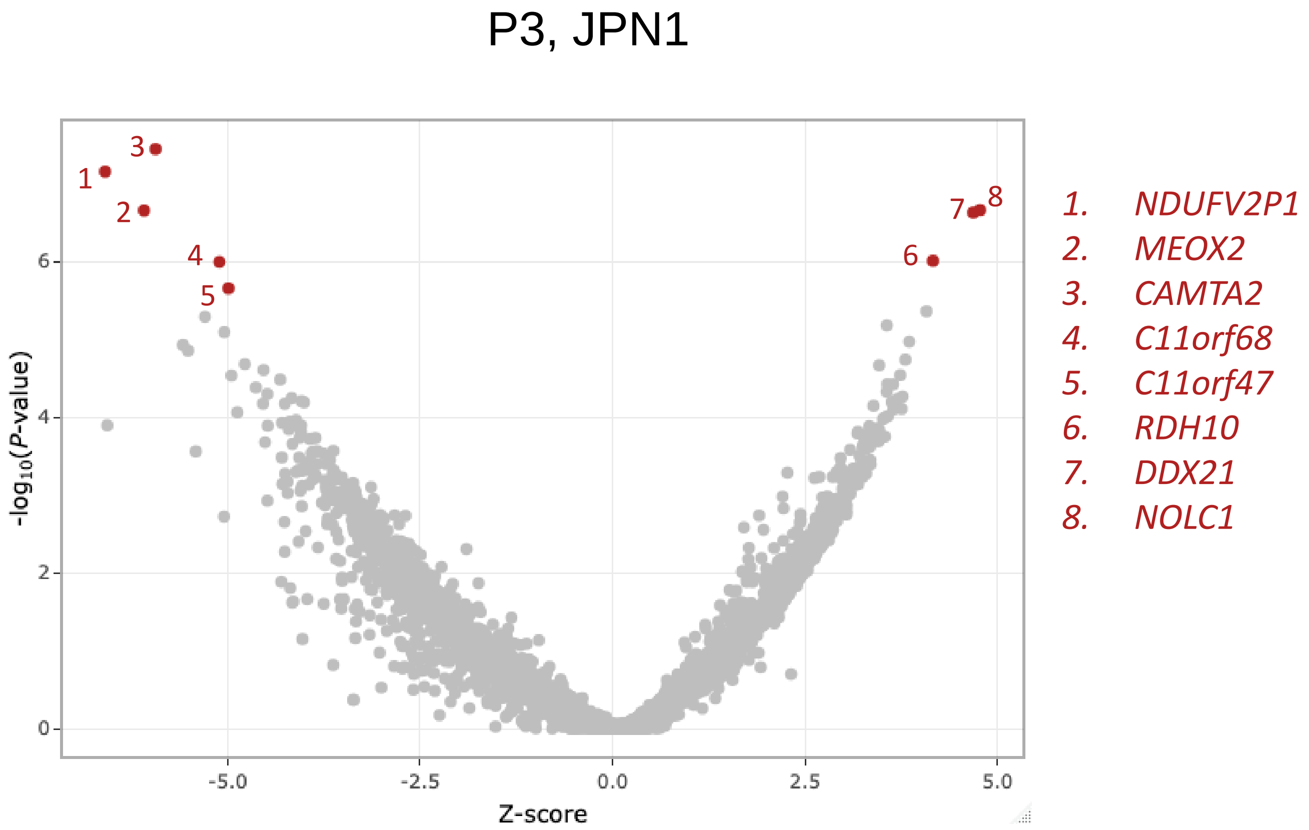


**Supplementary Figure 1. RNA-seq expression analysis for JPN1 using OUTRIDER (version 1.14.0).** Volcano plot of the RNA-seq data from patient P3 (JPN1) against 129 individuals showing gene expression significance values (−log10 *p* value) against Z-scores. Genes with significantly lower or higher expression (FDR ≤ 0.05) are marked in red dots (left to right: *NDUFV2P1*, *MEOX2*, *CAMTA2*, *C11orf68*, *C11orf47*, *RDH10*, *DDX21* and *NOLC1*). The expression outliers were detected using DROP v1.0.3., which leverages OUTRIDER (version 1.14.0). The cohort consisted of 131 skeletal muscle RNAseq from rare muscle disease patients and unaffected controls. The sample size is above the recommended cut-off (n > 50) for OUTRIDER to detect significant events with a false discovery rate (FDR) ≤ 0.05.

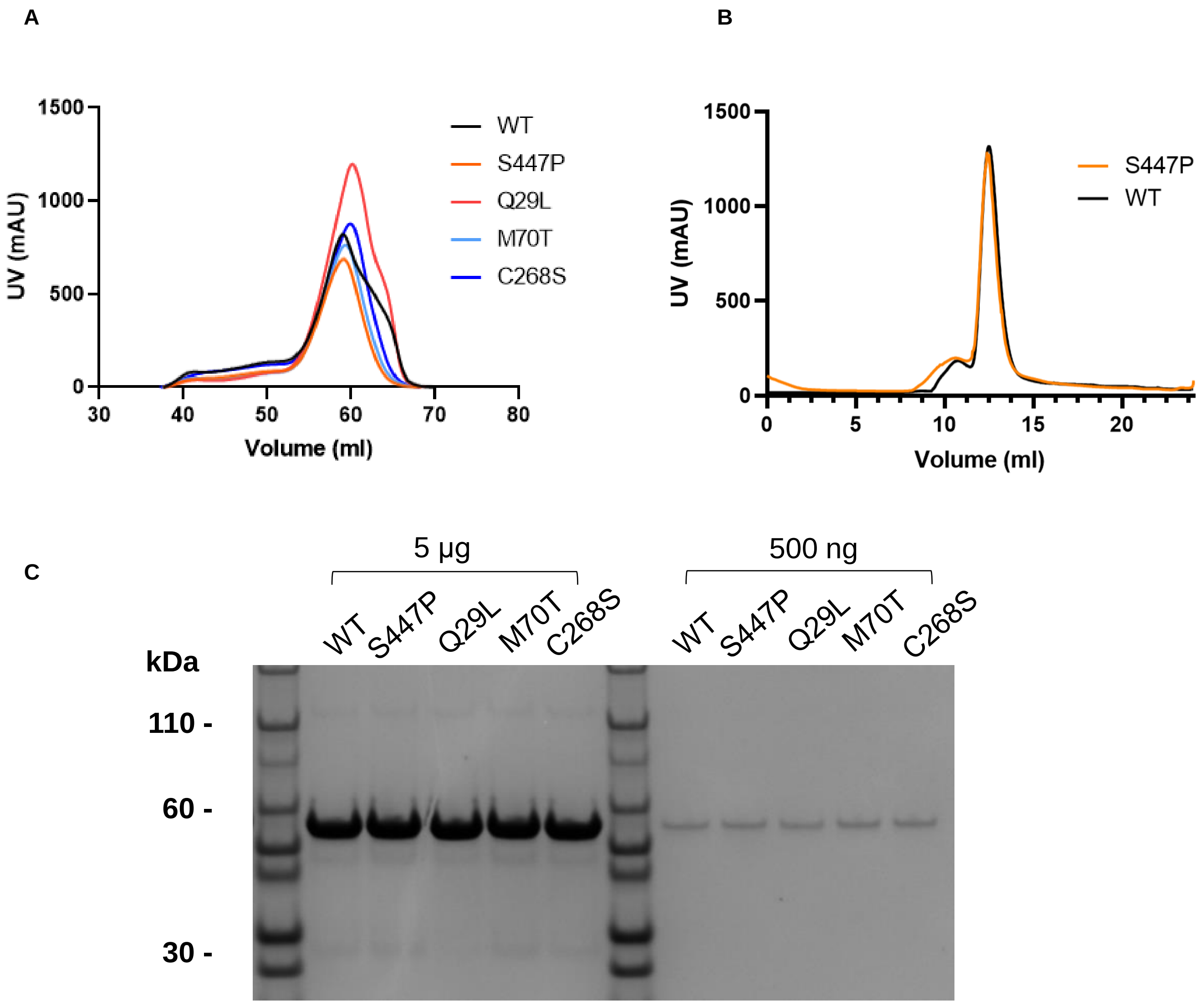


**Supplementary Figure 2. Evaluation of recombinant HMGCS1 purity and oligomerisation tendencies.** (A) Size exclusion chromatograms of recombinant wildtype (WT) and mutant (S447P, Q29L, M70T, C268S) HMGCS1 eluting at volumes (~60 mL) corresponding to the dimerised conformation. Proteins eluted by a XK16/600 Superdex 200 column (GE Healthcare). (B) Additional size exclusion chromatograms of HMGCS1WT and HMGCS1S447P using a 10/300 GL Superdex 200 column (GE Healthcare) indicates elution of both proteins at ~12 ml corresponding to the dimerised conformation. (C) SDS-PAGE evaluation of HMGCS1 protein eluting from the major size exclusion chromatography peaks in (A), confirming that all HMGCS1 were of the expected size (~57 kDa; monomer) and of satisfactory purity.

**A**

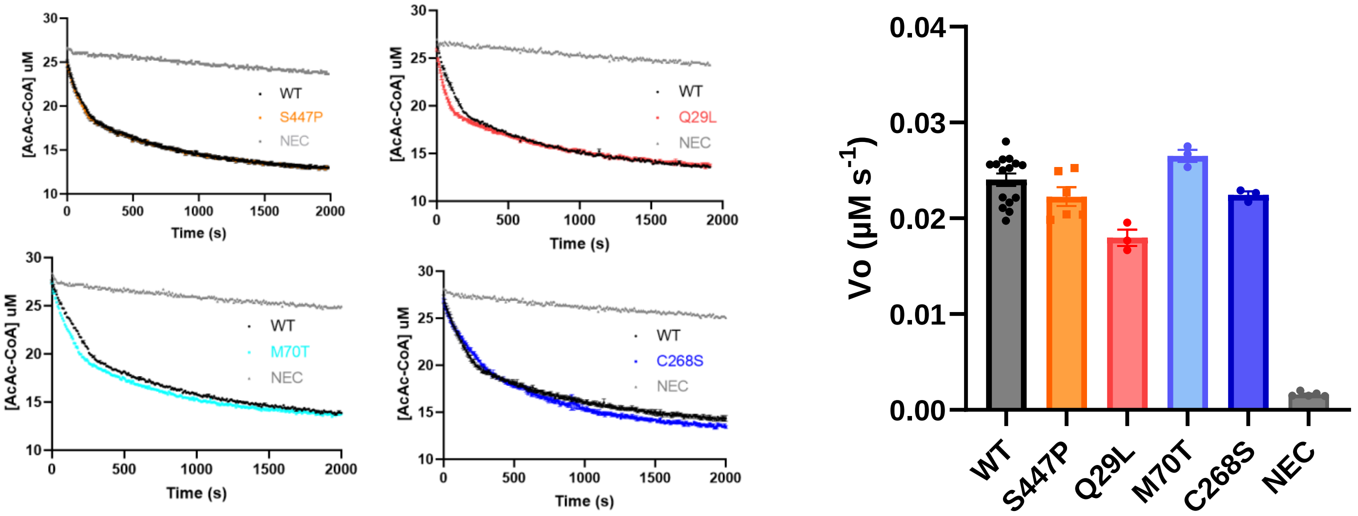


**B**

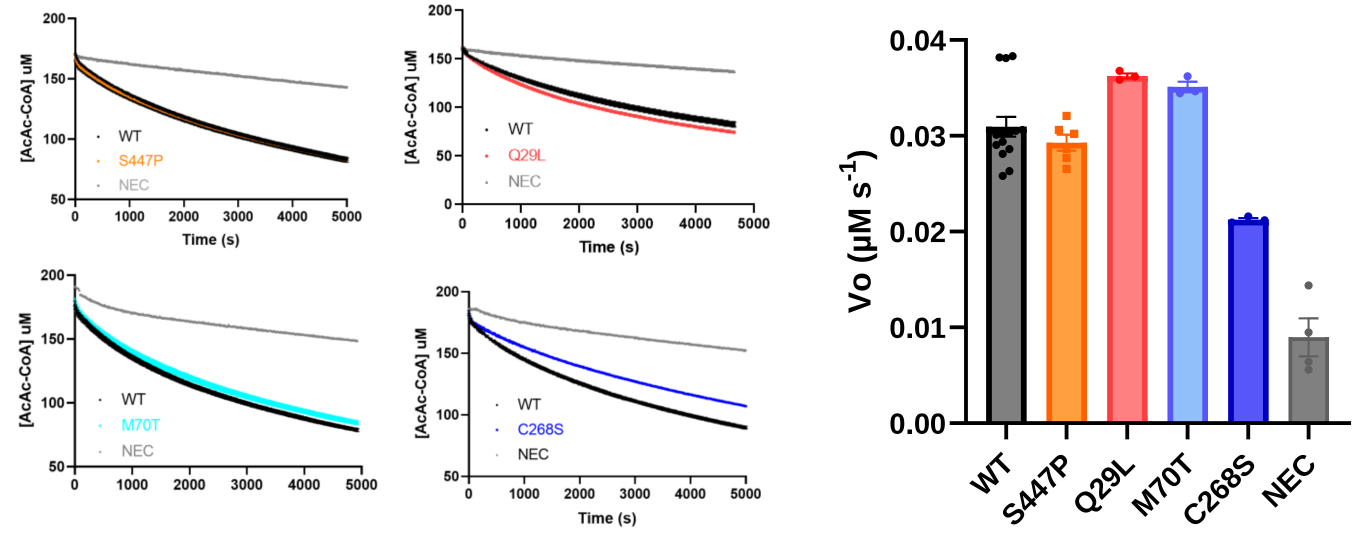


**Supplementary Figure 3 Enzyme assays of wildtype and mutant HMGCS1**. (A-B) Assays of wildtype (WT) and mutant (S447P, Q29L, M70T, C268S) HMGCS1 with (A) 25 µM AcAc-CoA and 200 µM Ac-CoA and (B) 200 µM AcAc-CoA and 200 µM Ac-CoA. The amount of AcAc-CoA consumed within the first 50 s of the reactions was taken as a measure of the initial velocity (V0). Assay buffer (100 mM Tris/HCl pH 8, 40 mM MgCl2) was used as a negative control (NEC).

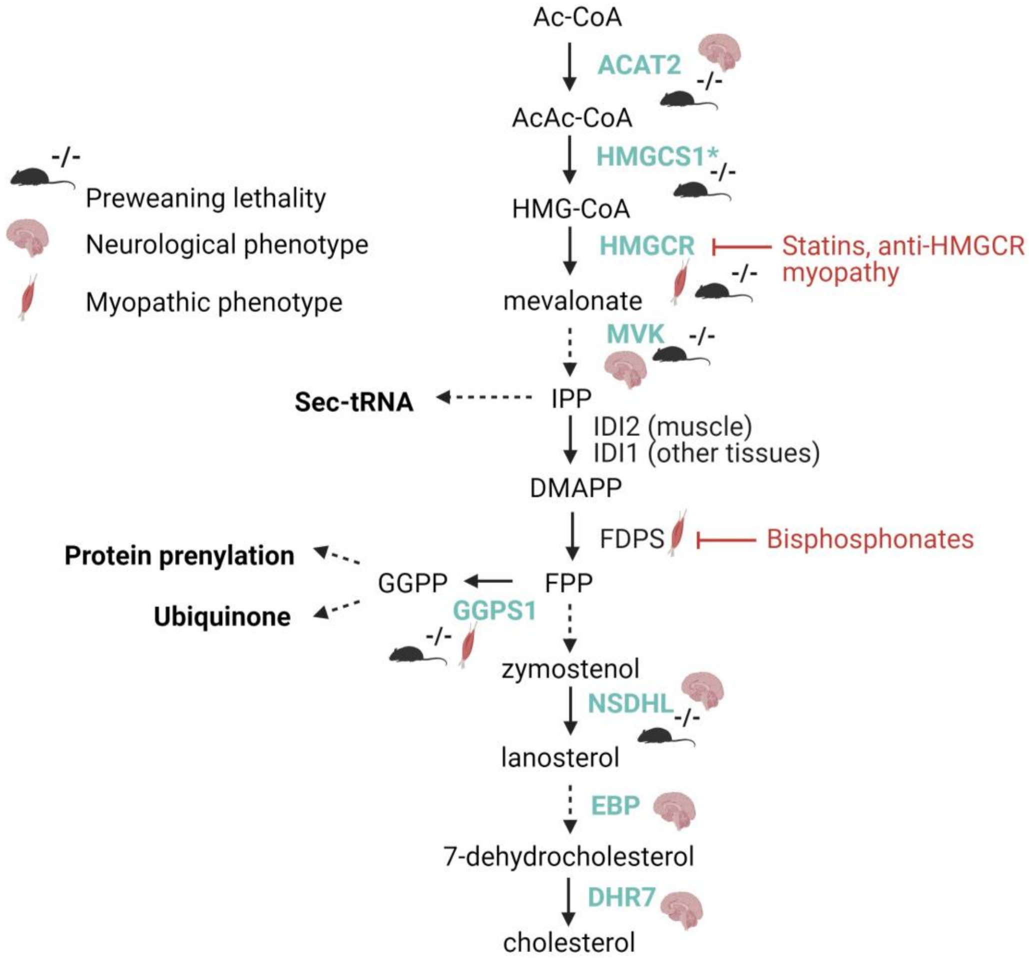


**Supplementary Figure 4 Diagrammatic summary of the mevalonate pathway.** Representative enzymes of the mevalonate pathway implicated in genetic disorders associated with neuromuscular phenotypes (blue). Mouse symbols represent genes associated with preweaning lethality when knocked out in mice. Muscle represents genes associated with myopathic manifestations. Brain represents genes associated with neurological manifestations. IDI2 is a skeletal muscle specific isopentenyl diphosphate isomerase isoform not yet associated with disease. Diagram produced by biorender.
